# MODELING CYSTIC FIBROSIS PATIENT PROGNOSIS: NOMOGRAMS TO PREDICT LUNG TRANSPLANTATION AND SURVIVAL PRIOR TO HIGHLY EFFECTIVE MODULATOR THERAPY

**DOI:** 10.1101/2023.09.25.23296112

**Authors:** Annalisa V. Piccorelli, Jerry A. Nick

## Abstract

**Background:** The duration of time a person with cystic fibrosis (pwCF) spends on the lung transplant waitlist is dependent on waitlist and post-transplant survival probabilities and can extend up to 2 years. Understanding the characteristics involved with lung transplant and survival prognoses may help guide decision making by the patient, the referring CF Center and the transplant team.

**Methods:** This study seeks to identify clinical predictors of lung transplant and survival of individuals with CF using 29,847 subjects from 2003-2014 entered in the Cystic Fibrosis Foundation Patient Registry (CFFPR).

**Results:** Predictors significant (p ≤ 0.05) in the final logistic regression model predicting probability of lung transplant/death were: FEV_1_ (% predicted), BMI, age of diagnosis, age, number of pulmonary exacerbations, race, sex, CF-related diabetes (CFRD), corticosteroid use, infections with *B. cepacia, P. aeruginosa, S. aureus*, MRSA, pancreatic enzyme use, insurance status, and consecutive ibuprofen use for at least 4 years. The final Cox regression model predicting time to lung transplant identified these predictors as significant FEV_1_ (% predicted), BMI, age of diagnosis, age, number of pulmonary exacerbations, race, sex, CF-related diabetes (CFRD), corticosteroid use, infections with *B. cepacia, P. aeruginosa, S. aureus*, MRSA, pancreatic enzyme use, and consecutive ibuprofen use for at least 4 years. The concordance indices were 0.89 and 0.92, respectively.

**Conclusions:** The models are translated into nomograms to simplify investigation of how various characteristics relate to lung transplant and survival prognosis individuals with CF not receiving highly effective CFTR modulator therapy.

## INTRODUCTION

The lung function complications due to cystic fibrosis (CF) lead to death in 63% of persons with CF, and as a result, persons with CF (pwCF) are often listed for a lung transplant [1]. Since late 2019 CF transmembrane-conductance regulator (CFTR) modulation is now standard of care for up to 90% of pwCF in the U.S. These small molecules directly modulate the activity and trafficking of the defective CFTR protein [2]. Of particular interest is “highly effective CFTR modulator therapy” (HEMT) which has resulted in exceptional improvement in mean lung function, body mass index, quality of life and rate of pulmonary exacerbations. HEMT commonly refers to use of ivacaftor in patients with CFTR gating mutations [4, 5], and the triple combination elexacaftor, tezacaftor and ivacaftor (E/T/I) in pwCF who are either homozygous or heterozygous for F508del mutation [3].

Prior to introduction of HEMT the waiting period on the lung transplant list could last 2 or more years and 23% of CF patients may die while on the list, understanding when to list them is a priority [6, 7] The Lung Allocation Score, which is used by the Organ Procurement and Transplantation Network to assure equitable organ allocation, takes survival probability while on the list and after transplantation into account, but is not tailored specifically to CF [8]. Following approval of E/T/I within the U.S. the 2020 CFFPR reported a notable reduction in the number of lung transplants performed [9].

People with CF are typically referred for lung transplant evaluation when their FEV_1_ (forced expiratory volume in 1 second) falls below 30 (% predicted), with female patients and younger patients often considered earlier [10]. Studies have identified other patient characteristics involved in CF lung transplant and survival, although the set of the predictors chosen differs depending on the study [6, 10, 11, 12, 13, 14, 15, 16, 17, 18, 19, 20, 21, 22, 23, 24, 25, 26, 27, 28, 29, 30, 31, 32, 33, 34, 35, 36, 37, 38, 39, 40]. Most of the studies indicate FEV_1_ % predicted (FEV1pp), age, number of pulmonary exacerbations, BMI, and sex are involved; however, the accuracy of these models often rely on many more.

Identifying the clinical characteristics involved in predicting when an individual will require a lung transplant may lead to a more personalized approach to transplant referral and listing. Additionally, these need to be communicated effectively from clinicians at CF Care Centers to pwCF and lung transplant centers [10]. The CFF consensus guidelines recommends “routine clinician-led efforts to discuss disease trajectory and treatment options, including lung transplantation” and “communication between the CF and lung transplant care teams at least every 6 months and with major clinical changes” [10].

The purpose of this study is to develop models to predict probability of lung transplant or death and time to lung transplant or death of CF patients in the United States prior to HEMT, and translate these into nomograms. The nomograms are meant to personalize estimations of survival and potentially ease communication between the CF clinician and patient and the lung transplant care team about the characteristics of the CF patient affecting both their probability of lung transplant or death and their probability of lung-transplant-free survival. Nkam et al. (2017) developed a nomogram predicting a patient’s 3-year risk of lung transplant or death given the patient’s characteristics based on data from the French CF Registry [30]. The current study aims to demonstrate the validity of this method using data pre-HEMT, and a step towards a current prediction of differing time periods of risk based on patient-specific characteristics to provide a more detailed understanding of a specific patient’s prognosis.

## METHODS

### Patients

This study was approved by the University of Wyoming Institutional Review Board. A subset of data from the CFF Patient Registry (CFFPR) was used in our study. The CFFPR collects data on CF patients who have been treated in CFF-accredited institutions and consented to be included in the CFFPR [41]. The CFFPR is estimated to include approximately 81-84% of persons with CF in the United States [41]. The data included 29,847 patients ages 6 to 40 years from from Jan. 1, 2003 to Dec. 31. 2014. This was a retrospective study of deidentified data and the authors did not have access to the patient identification. These age limits were included in this study, because pulmonary function tests cannot be measured reliably until age 6 and estimates of survival after age 40 are subject to survivor bias and severe left truncation [42]. The interval chosen pre-dates significant usage of HEMT in the US CF population.

### Predictors and Outcome variables

Lung transplant or death, whichever occurred first, was the event of interest for both the logistic and Cox regression models, described in more detail below. For the Cox model, time to event was set up as the minimum of the age of lung transplant or death, with age in the most recent review year used for censored observations. The data cleaning and summary measures calculations were conducted with SAS 9.4.

The predictor variables attempted are shown on Table 1 and are based on prior studies [6, 10, 11, 12, 13, 14, 15, 16, 17, 18, 19, 20, 21, 22, 23, 24, 25, 26, 27, 28, 29, 30, 31, 32, 33, 34, 35, 36, 37, 38, 39, 40]. The following demographic variables were considered: race, sex, F508del genotype, and age of diagnosis. Race was defined as whites v. other, because the majority (93.7%) were white. The following variables that were collected at encounter visits were also considered: weight, height, BMI, most recently documented age, best yearly FEV1 % predicted (FEV1pp), best yearly forced vital capacity % predicted (FVCpp), *Pseudomonas aeruginosa* (*P. aeruginosa*) infection, *Staphylococcus aureus* (*S. aureus*), methicillin-resistant *Staphylococcus aureus* (MRSA) infection, *Burkholderia cepacia* (*B. cepacia*) infection, CF related diabetes (CFRD), pancreatic enzyme use, high dose ibuprofen use for at least 4 consecutive years (high ibuprofen), corticosteroid therapy, insurance status, and number of pulmonary exacerbations treated with IV antibiotics (NumPulmExacerbation). All of the variables other than best yearly FEV1pp and FVCpp were based on annualized observations. Annualized indicates the data were converted from encounter visits; for quantitative variables, the highest measurement from each quarter was taken and then was averaged to create on measure for the year. Annualized observations were selected over encounter observations because there were less missing observations in the annualized dataset. Best yearly FEV1pp and FVCpp were calculated from encounter visits. Because there were more missing BMI values than height and weight values, BMI was calculated based on the formula: BMI = weight/height^2^. Annualized and best yearly observations were taken 1 year prior to the death/lung transplant (whichever was first) or at time of censoring.

**Table 1.** Summary of demographic, annualized, and best yearly pulmonary characteristics by event.

| Characteristic |  | Death/Transplant<br>5206 (17.4%) | Alive<br>24,641 (82.6%) |
| --- | --- | --- | --- |
| Demographic |  |  |  |
| Race* | White | 4979 (95.6%) | 23,033 (93.5%) |
| Sex* | Female | 2740 (52.6%) | 11,565 (46.9%) |
| F508del genotype<sup>m*</sup> | 1 | 2514 (48.3%) | 10,907 (44.3%) |
|  | 2 | 1595 (30.6%) | 9288 (37.7%) |
|  | 3 | 388 (7.5%) | 3595 (14.6%) |
| Age of diagnosis* | Mean (SD) | 2.3 (4.7) | 3.5 (6.6) |
| Annualized variables 1 year prior to event or at censoring |  |  |  |
| *B. cepacia*<sup>m*</sup> | Positive | 62 (1.2%) | 385 (1.6%) |
| CF related diabetes<sup>m*</sup> | 1 | 1924 (37.0%) | 17,237 (70.0%) |
|  | 2 | 375 (7.2%) | 2662 (10.8%) |
|  | 3 | 2907 (55.8%) | 4742 (19.2%) |
| On corticosteroid therapy<sup>m*</sup> | Positive | 1541 (29.6%) | 13,489 (54.7%) |
| *P. aeruginosa*<sup>m*</sup> | Positive | 2252 (43.3%) | 12,025 (48.8%) |
| *S. aureus*<sup>m*</sup> | Positive | 1516 (29.1%) | 16,603 (67.4%) |
| MRSA<sup>m*</sup> | Positive | 951 (18.3%) | 6331 (25.7%) |
| On pancreatic enzymes<sup>m*</sup> | Positive | 4889 (93.9%) | 20,985 (85.2%) |
| Insurance<sup>m*</sup> | Federal | 2677 (51.4%) | 8936 (36.3%) |
|  | Private | 2345 (45.0%) | 14,901 (60.5%) |
|  | Other | 27 (0.5%) | 209 (0.8%) |
|  | None | 43 (0.8%) | 328 (1.3%) |
| High Ibuprofen use for 4 consecutive years* | Positive | 23 (0.4%) | 482 (2.0%) |
| BMI<sup>m†*</sup> | Mean (SD) | 19.6 (3.5) | 20.9 (4.2) |
| Height<sup>m*</sup> | Mean (SD) | 162.4 (12.6) | 157.5 (18.8) |
| Weight* | Mean (SD) | 52.3 (13.5) | 54.0 (19.4) |
| Age* | Mean (SD) | 26.5 (7.7) | 20.6 (9.6) |
| NumPulmExacerbation<sup>‡*</sup> | Mean (SD) | 1.9 (2.0) | 0.7 (1.3) |
| Best yearly 1 year prior to event or at censoring |  |  |  |
| FEV1pp<sup>m*</sup> | Mean (SD) | 49.4 (27.2) | 83.0 (25.6) |
| FVCpp<sup>m*</sup> | Mean (SD) | 61.0 (23.3) | 92.5 (21.2) |

<sup>m</sup> Some observations were missing.
\*significant difference between groups (p-value<0.05), association between categorical predictor and response
†calculated from height and weight
‡number of pulmonary exacerbations treated with IV antibiotics.

The % missingness observed for these variables was as follows: F508del genotype (5.2%), *B. cepacia* (11.5%), corticosteroid therapy (13.0%), *P. aeruginosa* (11.5%), *S. aureus* (11.5%), MRSA (11.5%), pancreatic enzyme usage (1.5%), insurance (1.3%), FEV1pp (5.4%), FVCpp (5.3%), height (0.5%), weight (0.8%), age of diagnosis (0.1%), and BMI (1.4%). Missing observations were imputed using SAS PROC MI and the imputed dataset that resulted in the minimum standard error for the majority of the variables was used for model development.

### Model Development

The data was analyzed using the software R 4.0.3. Multiple logistic regression modeling was used to predict lung transplant or death and predict the probability of lung transplant or death. Cox regression models were used to identify significant predictors of time to lung transplant or death and probability of time to lung transplant or death. Full logistic and Cox regression models were fit including all independent variables of interest. Significance of predictors (p ≤ 0.05), Akaike information criterion (AIC), and concordance index were used to select the best subset of predictors for the final models. The Holm method was used to correct p-values for multiple testing [43]. The Cox proportional hazards assumption was checked using Schoenfeld residuals. Final models were used to generate nomograms of the probability of lung transplant or death and time to lung transplant-free survival.

### Model Validation

Leave one out cross-validation was used to determine the estimated prediction error of the logistic regression model using R function cv.glm with the number of resamples equal to the sample size of 29,847. Since there is not an R function for leave one out cross-validation with Cox regression modeling, bootstrapped resampling with 1000 repetitions was used to validate the Cox regression model with R function validate.

## RESULTS

### Patient Characteristics

The subset of the CFFPR eligible for this study was 29,847 CF patients. Descriptive statistics of the study sample are shown on Table 1. Death or lung transplant was observed in 5206 (17.4%) patients with the remaining 24,641 patients (82.6%) censored in 2014 or if missing in 2014, the time of most current review prior to 2014. Significant differences between the death/transplant and the alive groups were observed in the following characteristics: age of diagnosis, BMI, height, weight, age, FEV1pp, FVCpp, and number of pulmonary exacerbations treated with IV antibiotics (NumPulmExacerbation). Significant associations were observed between vital status and the following categorical variables: race, sex, F508del genotype, CFRD, corticosteroid therapy, *B. cepacia, P. aeruginos*a, *S. aureus*, MRSA, pancreatic enzymes therapy, insurance status, and high dose ibuprofen use for at least 4 consecutive years.

### Multiple Logistic Regression Model

The following characteristics were significant predictors of probability of lung transplant/death in the final logistic regression model (Table 2, Fig. 1): FEV1pp, BMI, age of diagnosis, age, NumPulmExacerbation, race, sex, CFRD, corticosteroid therapy, *B. cepacia, P. aeruginosa, S. aureus*, MRSA, pancreatic enzyme usage, insurance status, and consecutive high dose ibuprofen use for at least 4 years.

**Table 2.** Logistic regression results for probability of lung transplant/death using complete dataset.

|  | Estimate | Standard Error | p-value |
| --- | --- | --- | --- |
| Intercept | 1.90 | 0.183 | <0.0001 |
| Sex | -0.26 | 0.389 | <0.0001 |
| FEV1pp | -0.04 | 0.001 | <0.0001 |
| BMI | -0.13 | 0.007 | <0.0001 |
| NumPulmExacerbation<sup>‡</sup> | 0.23 | 0.012 | <0.0001 |
| Age | 0.06 | 0.003 | <0.0001 |
| Age of diagnosis | -0.03 | 0.004 | <0.0001 |
| Race | 0.43 | 0.089 | <0.0001 |
| *B. cepacia* | -0.90 | 0.153 | <0.0001 |
| *P. aeruginosa* | -0.94 | 0.045 | <0.0001 |
| *S. aureus* | 0.33 | 0.048 | <0.0001 |
| MRSA | -0.56 | 0.051 | <0.0001 |
| CFRD2* | -0.21 | 0.071 | 0.03122 |
| CFRD3* | 0.90 | 0.042 | <0.0001 |
| Ibuprofen use for $\leq 4$<br>consecutive years | -1.38 | 0.243 | <0.0001 |
| Insurance_none† | -0.61 | 0.189 | 0.01185 |
| Insurance_other† | -0.63 | 0.241 | 0.03122 |
| Insurance_private† | -0.37 | 0.040 | <0.0001 |
| Enzymes | 0.61 | 0.090 | <0.0001 |
| Corticosteroids | -0.92 | 0.042 | <0.0001 |

‡number of pulmonary exacerbations treated with IV antibiotics.
\*CFRD categories are indicator variables for CFRD2= Impaired Glucose Tolerance, and CFRD3 = CFRD with or without fasting hyperglycemia with the reference category CFRD1 = Normal Glucose Metabolism.
†The reference category for insurance status was federal.

**Figure 1.**
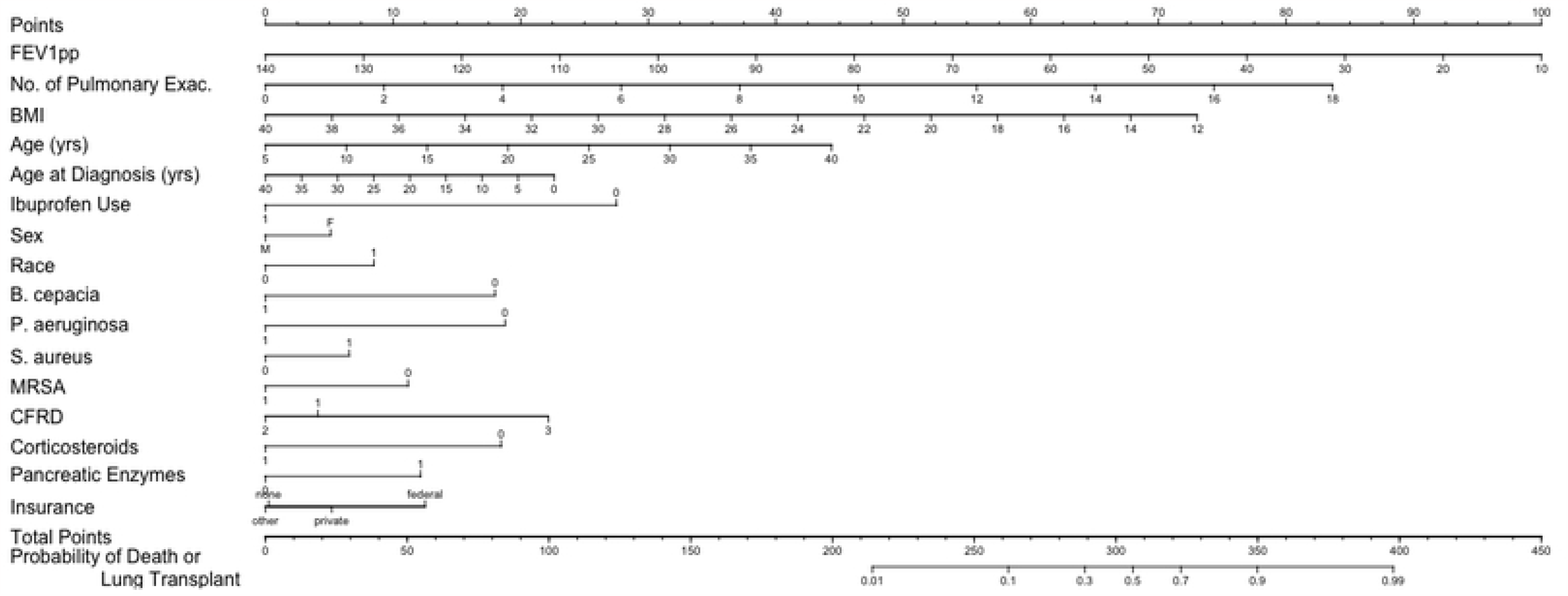
Nomogram for probability of lung transplant or death. For each characteristic, find the number of points by drawing a vertical line to the Points scale. Total the points for all the characteristics and draw a vertical line from the Total Points scale to get the probability of lung transplant or death.

### Cox Multiple Regression Model

The final Cox regression model of time to lung transplant/death identified the following significant predictors as: FEV1pp, BMI, age of diagnosis, age, NumPulmExacerbation, race, sex, CFRD, corticosteroid therapy, *B. cepacia, P. aeruginosa, S. aureus*, MRSA, pancreatic enzyme use, and consecutive high dose ibuprofen use for at least 4 years (Table 3, Fig. 2).

**Table 3.**
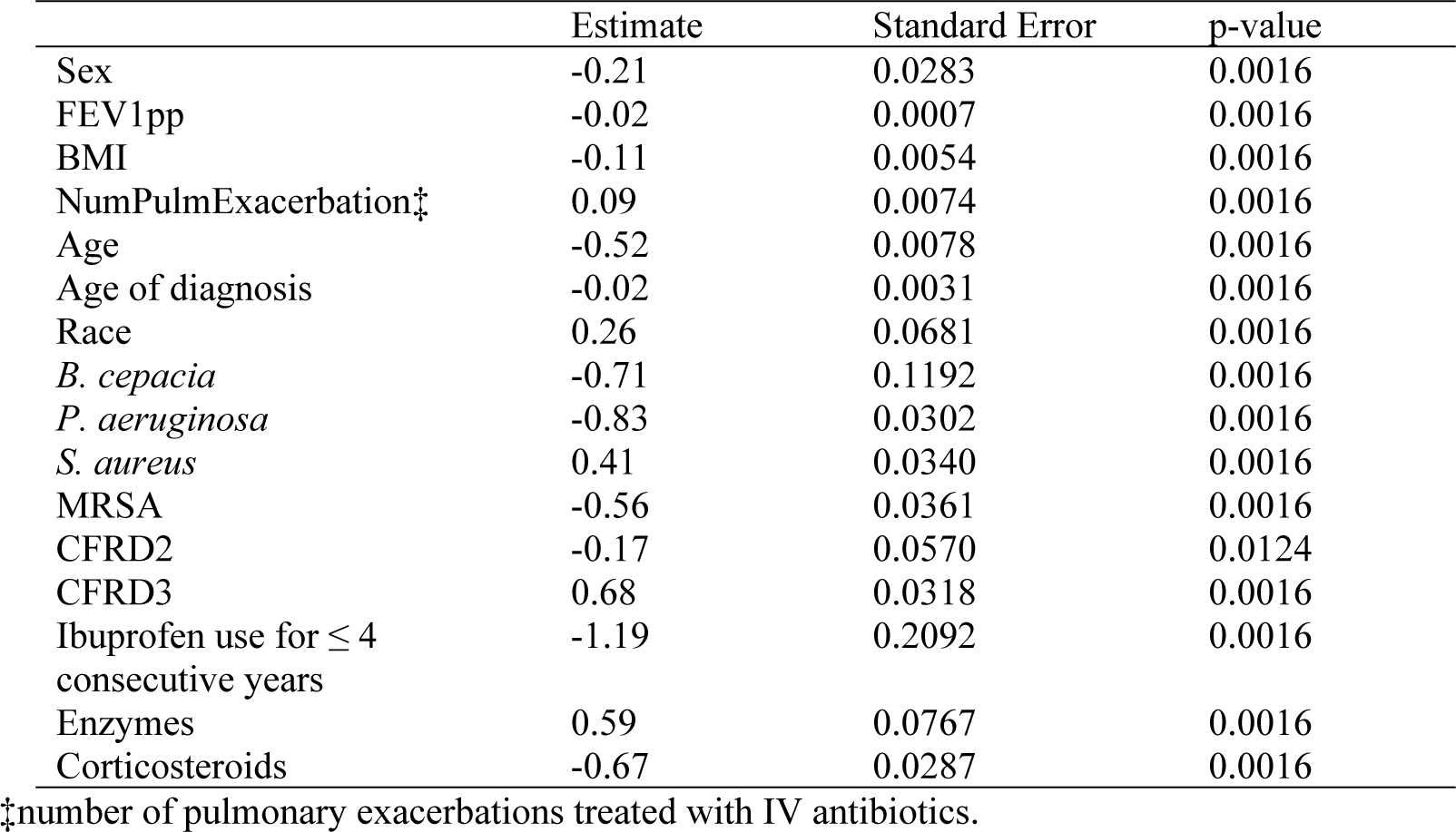
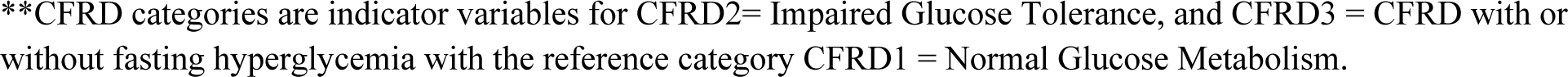
Cox regression results for probability of lung transplant/death prevention for 5 years using complete dataset.

|  | Estimate | Standard Error | p-value |
| --- | --- | --- | --- |
| Sex | -0.21 | 0.0283 | 0.0016 |
| FEV1pp | -0.02 | 0.0007 | 0.0016 |
| BMI | -0.11 | 0.0054 | 0.0016 |
| NumPulmExacerbation‡ | 0.09 | 0.0074 | 0.0016 |
| Age | -0.52 | 0.0078 | 0.0016 |
| Age of diagnosis | -0.02 | 0.0031 | 0.0016 |
| Race | 0.26 | 0.0681 | 0.0016 |
| *B. cepacia* | -0.71 | 0.1192 | 0.0016 |
| *P. aeruginosa* | -0.83 | 0.0302 | 0.0016 |
| *S. aureus* | 0.41 | 0.0340 | 0.0016 |
| MRSA | -0.56 | 0.0361 | 0.0016 |
| CFRD2 | -0.17 | 0.0570 | 0.0124 |
| CFRD3 | 0.68 | 0.0318 | 0.0016 |
| Ibuprofen use for $\leq 4$<br>consecutive years | -1.19 | 0.2092 | 0.0016 |
| Enzymes | 0.59 | 0.0767 | 0.0016 |
| Corticosteroids | -0.67 | 0.0287 | 0.0016 |

‡number of pulmonary exacerbations treated with IV antibiotics.
\*\*CFRD categories are indicator variables for CFRD2= Impaired Glucose Tolerance, and CFRD3 = CFRD with or without fasting hyperglycemia with the reference category CFRD1 = Normal Glucose Metabolism.

**Figure 2.**
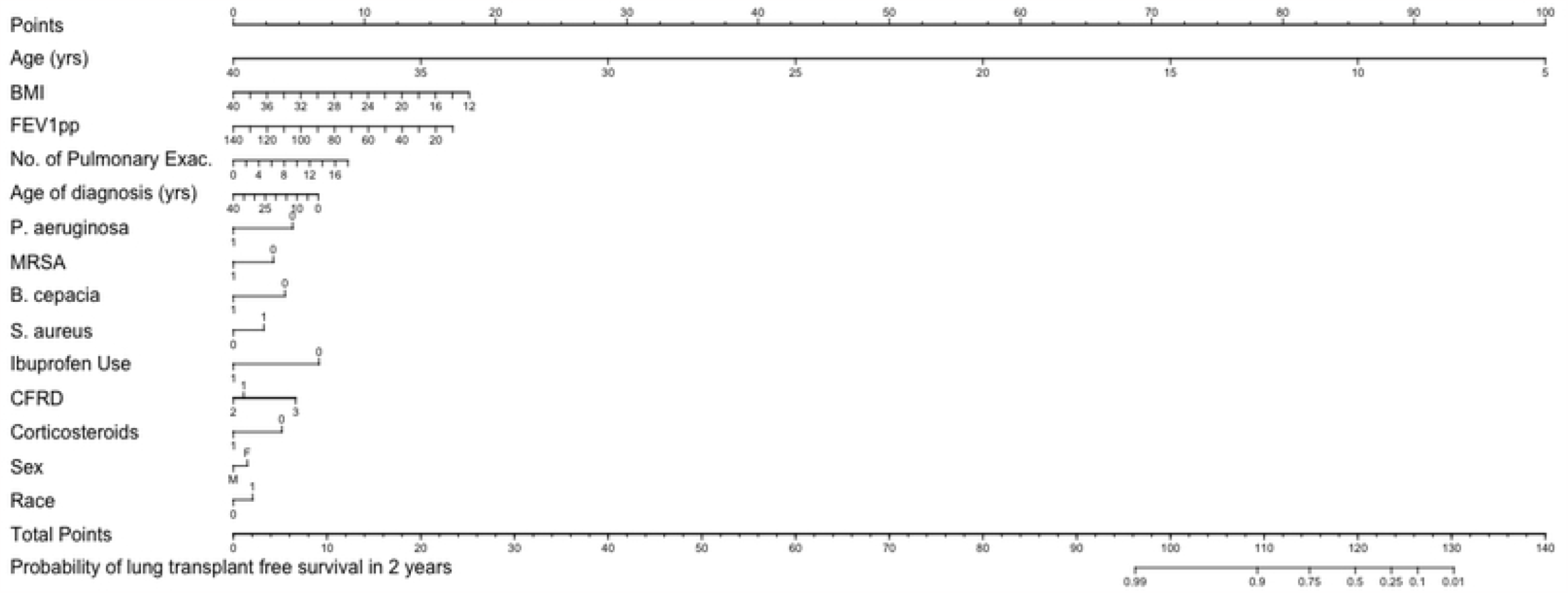
Nomogram for probability of lung transplant or death free survival for 2 years. Notes about variables: No. of Pulmonary Exac: number of pulmonary exacerbations treated with IV antibiotics in most recent year, Race: 0 = non-white, 1 = white; Sex: M = Male, F = Female; CFRD: 1 = Normal Glucose Metabolism, 2 = Impaired Glucose Tolerance, 3 = CFRD with or without fasting hyperglycemia; for variables, *B. cepacia, P. aeruginosa, S. aureus*, and MRSA: 0 = negative and 1 = positive; for variables, Corticosteroid Therapy (Corticosteroids) and Pancreatic enzyme usage (Pancreatic Enzymes): 0 = No, 1 =Yes; Insurance: F = Federal, P = Private, O = Other, and N = None; Ibuprofen use: use of ibuprofen use for at least 4 consecutive years.

### Model Validation

The validation of the model was measured based concordance index measuring the proportion of the predictions of the models to predict lung transplant/death in patients that experienced lung transplant/death. The multiple logistic regression model had a concordance index of 0.89 and an estimated prediction error of 0.09 from leave one out cross-validation. The Cox multiple regression model performed similarly as well with a concordance index of 0.92 based on the bootstrap resampling validation.

## DISCUSSION

This study developed models that were able to accurately predict probability of the lung transplant/death and time to lung transplant/death. The logistic and Cox regression models were internally validated with accuracy of prediction at 89% and 92%, respectively. The logistic regression model predicting probability of lung transplant/death identified FEV1pp, BMI, age of diagnosis, age, NumPulmExacerbation, race, sex, CFRD, corticosteroid therapy, *B. cepacia, P. aeruginosa, S. aureus*, MRSA, pancreatic enzyme usage, insurance status, and consecutive high dose ibuprofen use for at least 4 years. Similarly the following characteristics were identified by the Cox regression modeling time to lung transplant/death: FEV1pp, BMI, age of diagnosis, age, NumPulmExacerbation, race, sex, CFRD, corticosteroid therapy, *B. cepacia, P. aeruginosa, S. aureus*, MRSA, pancreatic enzyme use, and consecutive high dose ibuprofen use for at least 4 years. Significant differences between and associations with vital status were observed in all the characteristics identified by the logistic and Cox regression models.

The logistic and Cox regression models were translated into nomograms. These nomograms are intended to ease the communication between CF clinicians and people with CF, and the lung transplant care center. The characteristics included in the nomograms are measured at most encounter visits (see Methods section for more detail), thus, the CF clinician can routinely calculate the CF patient’s probability of lung transplant and probability of lung transplant-survival in 2 years and 5 years. This may enable the CF clinician and the individual with CF to stay on track with a timely lung transplant referral, to avoid complications and potential barriers to listing associated with late referrals [10]. This is intended help to address morality prior to listing, which is reported as high as 50% in a recent study from France [44]. The additional benefit of the modeling time to lung-transplant free survival with a Cox regression is the ability to predict probability of lung transplant/death in different time-points allowing investigators to choose time-points that are meaningful to a specific person with CF.

Nkam et al. (2017) also developed a nomogram to predict probability of lung transplant. Their model predicted probability of lung transplant in 3 years based on a smaller sample (n=2096) of the French CF Registry with only 3 years of data (2010-13) [30]. Their model included the following categorical predictors: *B. cepacia*, hospitalization (yes/no), oral corticosteroids, long-term oxygen therapy, and non-invasive ventilation, and categorical versions of FEV1pp (≥ 60, [30-60], <30) and BMI (≥ 18.5, [16-18.5], <16). Although the current study’s models are more complex in terms are the number of predictors, they can be more specific to each patient given the models use non-categorical versions of the quantitative predictors and can predict probability of lung transplant-free survival at different time-points.

This study was limited by the variables available in the CFFPR. Characteristics suggested to exacerbate lung function and affect survival by other papers, such as infection with *Stenotrophomonas maltophilia*, 6 minute walking test, hypercarbia, and hypoxemia, were not available in the CFFPR [10, 45, 46].

Although nomograms allow for easy communication, there are limitations involved with applying them to predicting probability of and timing of lung transplant/death. Nomograms do not accommodate time-varying covariates, which could have been useful with conditions that change prognosis after they are present, e.g., *B. cepacia*. Given the necessity of nomograms to allow for clear, easy-to-implement-and-interpret, although other models suggested slope of FEV1 and interactions between pulmonary exacerbations and FEV1pp as predictors, adding these into the model would have made the nomogram more complex to interpret [25, 38]. Finally, the current nomograms were developed using data that generally predated widespread use of highly effective CFTR modulator therapy (HEMT) in the United States CF population. CFTR modulators improve outcomes for a majority of individuals with CF [2, 3, 5] and may deliver a sustained benefit over time [47]. Clinical trials excluded patients with advanced lung disease, thus there is little available data to guide the degree of disease modification in the period included within this study. It is now apparent that most individuals receiving HEMT will experience slower progression towards transplant or death in this cohort, but current CFF-sponsored guidelines for lung transplant referral suggest that transplant referral not be delayed based on use of CFTR modulators [2, 3, 5, 47]. Going forward, these findings can serve as a comparison for future cohorts who are on HEMT.

We have developed and internally validated nomograms to predict probability of lung transplant/death and probability of lung transplant-free survival in 2 year and 5 years. The nomograms are user-friendly and will facilitate further investigation into need for transplant and survival in people with CF with advanced lung disease.

## Data Availability

The data underlying the results presented in the study are available from the Cystic Fibrosis National Patient Registry (https://www.cff.org/medical-professionals/patient-registry).

## Acknowledgements

The authors would like to thank the Cystic Fibrosis Foundation for the use of the CF Foundation Patient Registry data to conduct this study. Additionally, we would like to thank the patients, care providers, and clinic coordinators at CF Centers throughout the United States for their contributions to the CF Foundation Patient Registry.

## References

[1] U.S. Cystic Fibrosis Foundation. Patient registry (2018) 2017 Annual Data Report. Bethesda, MD.

[2] Heijerman, H.G.M., McKone, E.F., Downey, D.G., Van Braeckel, E., Rowe, S.M., Tullis, E., et al. (2019). Efficacy and safety of the elexacaftor plus texacaftor plus ivacaftor combination regimen in people with cystic fibrosis homozygous for the F508del mutation: a double-blind, randomised, phase 3 trial. Lancet 394: 1940–1948. 10.1016/S0140-6736(19)32597-8.

[3] Middleton, P.G., Mall, M.A., Dievinek, P., Lands, L.C., McKone, E.F., Polineni, D., et al. (2019). Elexacaftor-Tezacaftor-Ivacaftor for Cystic Fibrosis with a Single Phe508del Allele. N Engl J Med. 381(19):1809–1819. 10.1056/NEJMoa1908639.

[4] Davies, J.C., Wainwright, C.E., Canny, G.J., Chilvers, M.A., Howenstine, M.S., Munck, A., et al. (2013). Efficacy and safety of ivacaftor in patients aged 6 to 11 years with cystic fibrosis with a G551D mutation. Am J Respir Crit Care Med 187: 1219–1225. 10.1164/rccm.201301-0153OC.

[5] Ramsey, B.W., Davies, J., McElvaney, N.G., Tullis, E., Bell, S.C., Dřevínek, P., et al. (2011). A CFTR Potentiator in Patients with Cystic Fibrosis and the G551 Mutation. N Engl J Med. 365(18):1663–72. 10.1056/NEJMoa1105185.

[6] Belkin, R.A., Henig, N.R., Singer, L.G., Chaparro, C., Rubenstein, R.C., Xie, S.X., et al. (2006). Risk factors for death of patients with cystic fibrosis awaiting lung transplantation. Am. J. Respir. Crit. Care Med. 173: 659–666. 10.1164/rccm.200410-1369OC.

[7] Health Resources and Services Administration. OPTN (Organ Procurement and Transplantation)/SRTR (Scientific Registry of Transplant Recipients) 2017 Annual Data Report: Lung. (2018). https://srtr.transplant.hrsa.gov/annual_reports/2017/Lung.aspx. 2017

[8] Egan, T.M., Murray, S., Bustami, R.T., Shearon, T.H., McCullough, K.P., Edwards, L.B., et al. (2006). Development of the New Lung Allocation System in the United States. American Journal of Transplantation. 6(Part 2): 1212–1227. 10.1111/j.1600-6143.2006.01276.x.

[9] U.S. Cystic Fibrosis Foundation. Patient Registry (2021) 2020 Annual Data Report. Bethesda, MD

[10] Ramos, K.J., Smith, P.J., McKone, E.F., Pilewski, J.M., Lucy, A., Hempstead, S.E., et al. (2019). Lung transplant referral for individuals with cystic fibrosis: Cystic Fibrosis Foundation consensus guidelines. J Cyst Fibros. 18: 312–333. 10.1016/j.jcf.2019.03.002.

[11] Aaron, S.D., Stephenson, A.L, Cameron, D.W., and Whitmore, G.A. (2015). A statistical model to predict one-year risk of death in patients with cystic fibrosis. J Clin Epi. 68: 1336–1345. 10.1016/j.jclinepi.2014.12.010.

[12] Augarten, A., Akons, H., Aviram, M., Bentur, L., Picard, E., Rivlin, J., et al. (2001). Prediction of mortality and timing of referral for lung transplantation in cystic fibrosis patients. Pediatr Transplant. 5: 339–342. 10.1034/j.1399-3046.2001.00019.x.

[13] Corey, M., Farewell, V. (1995). Determinants of Mortality from Cystic Fibrosis in Canada, 1970-1989. American Journal of Epidemiology. 143(10): 1007–17. 10.1093/oxfordjournals.aje.a008664.

[14] Dasenbrook, E.C., Merlo, C.A., Diener-West, M., Lechtzin, N., Boyle, M.P. (2008). Persistent methicillin-resistant Staphylococcus aureus and rate of FEV_1_ decline in cystic fibrosis. Am. J. Respir. Crit. Care Med. 178: 814–821. 10.1164/rccm.200802-327OC.

[15] de Boer K., Vandemheen, K.L., Tullis, E., Doucettte, S., Fergusson, D., Freitag, A., et al. (2011). Exacerbation frequency and clinical outcomes in adult patients with cystic fibrosis. Thorax. 66: 680–685. 10.1136/thx.2011.161117.

[16] Ellaffi, M., Vinsonneau, C., Coste, J., Hubert, D., Burgel,P-R., Dhainaut, J-F., et al. (2005). One-year outcome after severe pulmonary exacerbation in adults with cystic fibrosis. Am. J. Respir. Crit. Care Med. 171: 158–64. 10.1164.rccm.200405-667OC.

[17] Emerson, J., Rosenfeld, M., McNamara, S., Ramsey, B., Gibson, R.L., (2002). Pseudomonas Aeruginosa and other predictors of mortality and morbidity in young children with cystic fibrosis. Pediatric Pulmonology. 34: 91–100. 10.1002/ppul.10127.

[18] George, P.M., Banya, W., Pareek., N, Bilton, D., Cullinan, P., Hodson, M.E., et al. (2011). Improved survival at low lung function in cystic fibrosis: cohort study from 1990-2007. BMJ. 342: d1008 10.1136/bmj.1008.

[19] Harness-Brumley, C.L., Elliott, A.C., Rosenbluth, D.B., Raghavan, D., Jain, R. (2014). Gender differences in outcomes of patients with cystic fibrosis. Journal of Women’s Health. 23: 1012–1020. 10.1089/jwh.2014.4985.

[20] Hayllar, K.M., Williams, S.C, Wise, A.E., Pouria, S., Lombard, M., Hudson, M.E., et al. (1997). A prognostic model for the prediction of survival in cystic fibrosis. Thorax. 52:313–317. 10.1136/thx.52.4.313.

[21] Lai, H.J., Cheng, Y., Cho, H., Kosorok, M.R., Farrell, P.M. (2004). Association between initial disease presentation, lung disease outcomes, and survival in patients with cystic fibrosis. Am. J. Epidemiol. 159: 537–546. 10.1093/aje/kwh083.

[22] Konstan, M.W., Byard, P.J., Hoppel, C.L., Davis, P.B. (1995). Effect of high-dose ibuprofen in patients with cystic fibrosis. New England Journal of Medicine. 332 (13): 848–854. 10.1056/NEJM199503303321303.

[23] Konstan, M.W., Butler, S.M., Wohl, M.E.B., Stoddard, M., Matousek, R., Wagener, J.S. (2003). Growth and nutritional indexes in early life predictor pulmonary function in cystic fibrosis. The Journal of Pediatrics. 142: 624–630. 10.1067/mpd.2003.152.

[24] Konstan, M.W., Schluchter, M.D., Xue, W., Davis, P.B. (2007). Clinical use of ibuprofen is associated with slower FEV_1_ decline in children with cystic fibrosis. Am. J. Respir. Crit. Care Med. 176: 1084–1089. rccm.200702-181OC.

[25] Liou, T.G., Adler, F.R., FitzSimmons, S.C., Cahill, B.C., Hibbs, J.R., Marshall, B.C. (2001b). Predictive 5-year survivorship model of cystic fibrosis. Am J Epidemiol. 153: 345–352. aje/153.4.345.

[26] Liou, T.G., Adler, F.R., Cox, D.R., Cahill, B.C., (2007). Lung transplantation and survival in children with cystic fibrosis. N Engl J Med. 357 (21): 2143–2152. 10.1056/NEJMx080021.

[27] Mayer-Hamblett, N., Rosenfeld, M., Emerson, J., Goss, C.H., Aitken, M.L. (2002). Developing cystic fibrosis lung transplant referral criteria using predictors of 2-year mortality. Am. J. Respir. Crit. Care Med. 166: 1550–1555. 10.1164/rccm.200202-087OC.

[28] Milla, C.E., Billings, J., Moran, A. (2005). Diabetes is associated with dramatically decreased survival in female but not male subjects with cystic fibrosis. Diabetes Care. 28: 2141–2144. 10.2337/diacare.28.9.2141.

[29] Nick, J.A., Chacon, C.S., Brayshaw, S.J., Jones, M.C., Barboa, C.M., St. Clair, C.G., et al. (2010). Effects of gender and age at diagnosis on disease progression in longterm survivors of cystic fibrosis. Am. J. Respir. Crit. Care Med. 182: 614–626. 10.1164/rccm.201001-0092OC.

[30] Nkam, L., Lambert, J., Latouche, A., Bellis, G., Burgel, P.R., Hocine, M.N. (2017). A 3-year prognostic score for adults with cystic fibrosis. J Cyst Fibros. 16: 702–708. 10.1016/j.jcf.2017.03.004.

[31] Orens, J.B., Estenne, M., Arcasory, S., Conte, J.V., Corris, P., Egan, J.J., et al. (2006). International Guidelines for the Selection of Lung Transplant Candidates: 2006 Update – A Consensus Report From the Pulmonary Scientific Council of the International Society for Heart and Lung Transplantation. J Heart Lung Transplant. 25(7): 745–755. 10.1016/healun.2006.03.011.

[32] Ramos, K.J., Quon, B.S., Heltshe, S.L., Mayer-Hambeltt, N., Lease, E.D., Aitken, M.L., et al. (2017). Heterogeneity in Survival in Adult Patients with FEV1 < 30% of Predicted in the United States. Chest. 151(6):1320–1328. 10.1016/j.chest.2017.01.019.

[33] Rodman, D.M., Polis, J.M., Heltshe, S.L., Sontag, M.K., Chacon, C., Rodman, R.V., et al. (2005). Late Diagnosis defines a unique population of Long-term survivors of cystic fibrosis. Am. J. Respir. Crit. Care Med. 171: 621–626. 10.1164/rccm.200403-404OC.

[34] Rosenbluth, D.B., Wilson, K., Ferkol, T., Schuster, D.P. (2004). Lung function decline in cystic fibrosis patients and timing for lung transplantation referral. Chest. 126: 412–9. 10.1378/chest.126.2.412.

[35] Rosenfeld, M., Davis., R., FitzSimmons, S., Pepe, M., Ramsey, B. (1997). Gender Gap in Cystic Fibrosis Mortality. Am J Epidemiol. 145: 794–803. 10.1093/oxfordjournals.aje.a009172.

[36] Sharma, R., Florea, V.G., Bolger, A.P., Doehner, W., Florea, N.D., Coats, A.J., et al. (2001). Wasting as an independent predictor of mortality in patients with cystic fibrosis. Thorax. 56: 746–750. 10.1136/thorax.56.10.746.

[37] Steinkamp, G., Wiedemann, B. (2002). Relationship between nutritional status and lung function in cystic fibrosis: cross-sectional and longitudinal analyses from the German CF quality (CFQA) project. Thorax. 57: 596–601. 10.1136/thorax.57.7.596.

[38] Stephenson, A.L., Tom, M., Berthiaume, Y., Singer, L.G., Aaron, S.D., Whitmore, G.A., et al. (2015). A contemporary survival analysis of individuals with cystic fibrosis: a cohort study. Eur Respir J. 45: 670–679. 10.1183/09031936.00119714.

[39] Vizza, C.D., Yusen, R.D., Lynch, J.P., Fedele, F., Patterson, G.A., Trulock, E.P. (2000). Outcome of patients with cystic fibrosis awaiting lung transplantation. Am. J. Respir. Crit. Care Med. 162: 819–825. ajrccm.162.3.9910102.

[40] Waters, V., Stanojevic, S., Atenafu, E.G., Lu, A., Yau, Y., Tullis, E., et al. (2012a). Effect of pulmonary exacerbations on long-term lung decline in cystic fibrosis. Eur Respir J. 40: 61–66. 10.1183/09031936.00159111.

[41] Knapp, E.A., Fink, A.K., Goss, C.H., Sewall, A., Ostrenga, J., Dowd, C., et al. (2016). The Cystic Fibrosis Foundation Patient Registry Design and Methods of a National Observational Disease Registry. Ann Am Thorac Soc. 13(7):1174–1179. 10.1513/AnnalsATS.201511-781OC.

[42] Schluchter, M.D., Konstan, M.W., and Davis, P.B. (2002). Jointly modeling the relationship between survival and pulmonary function in cystic fibrosis patients. Statistics in Medicine. 21: 1271–1287. 10.1002/sim.1104.

[43] Holm, S. (1979). A simple sequentially rejective multiple test procedure. Scandinavian Journal of Statistics, 6, 65–70. https://www.jstor.org/stable/4615733.

[44] Martin, C., Hamard, C., Kanaan, R., Boussaud, V., Grenet, D., Abély, M., et al. (2016). Causes of death in French cystic fibrosis patients: the need for improvement in transplantation referral strategies! J Cyst Fibros. 15(2):204–212. 10.1016/j.jcf.2015.09.002.

[45] Waters, V., Yau, Y., Prasad, S. Lu, A., Atenafu, E., Crandall, I. et al. (2012b). Stenotrophomonas maltophilia in Cystic Fibrosis Serologic Response and Effect on Lung Disease. Am. J. Respir. Crit. Care Med. 183: 635–640. 10.1164/rccm.201009-1392OC.

[46] Waters, V., Atenafu, E.G., Salazar, J.G., Lu, A., Yau, Y., Matukas, L. et al. (2013). Chronic Stenotrophomonas maltophilia infection and mortality or lung transplantation in cystic fibrosis patients. J Cyst Fibros. 12: 482–486. 10.1016/j.jcf.2011.07.008.

[47] Sawicki, G.S., McKone, E.F., Pasta, D.J., Millar, S.J., Wagener, J.S., Johnson, C.A., et al. (2015). Sustained Benefit from Ivacaftor Demonstrated by Combining Clinical Trial and Cystic Fibrosis Patient Registry Data. Am. J. Respir. Crit. Care Med. 192(7):836–42. 10.1164/rccm.201503-0578OC.

